# Application of Large Language Models (LLM) for Automatic Classification of Work Accident Text Data: Verification of Accuracy and Practicality

**DOI:** 10.1101/2025.10.02.25337141

**Authors:** Hajime Ando, Ryutaro Matsugaki, Sakumi Yamakawa, Akira Ogami

## Abstract

**Background:** Falls are the most frequent type of occupational accident, making the development of effective countermeasures an urgent issue. Traditional accident analysis relies on manual classification of text data by experts, a process that is both time-consuming and labor-intensive. Large Language Models (LLMs) offer the potential to significantly streamline this analysis process without the need for task-specific pre-training.

**Objective:** This study aims to automatically classify text data from occupational accidents using LLMs and to verify the accuracy and practicality of this approach.

**Methods:** The analysis targeted 2,619 fall-related injury cases in the health/hygiene and retail sectors, extracted from the 2021 Survey on Industrial Accidents database. The results of manual classification performed by experts in a previous study were used as the “ground truth.” These were compared against the automatic classification results from four different LLMs (GPT-4.1, GPT-4.1 mini, GPT-4o mini, and o4-mini). Evaluation metrics included accuracy, precision, recall, F1-score, and Cohen’s kappa coefficient. The processing was conducted using OpenAI’s Batch API, with processing time and costs also being measured.

**Results:** Newer generation models demonstrated a high rate of agreement with expert classifications across most categories, with the exception of “causal substance,” generally achieving a Cohen’s kappa coefficient above 0.7. For the “accident location (indoor/outdoor)” category, the accuracy reached over 91%. Even for “causal substance,” the category with the lowest accuracy, the reasoning model o4-mini achieved a kappa coefficient of 0.662. In terms of practicality, even when using the highest-performing model (o4-mini), the entire dataset was processed in approximately 90 minutes at a cost of about $11, demonstrating high cost-performance.

**Conclusion:** This study demonstrates that LLMs can classify occupational accident text data with an accuracy comparable to manual expert analysis, but at a lower cost and higher speed. This method is expected to facilitate large-scale accident analysis, which has been challenging in the past, and contribute to the rapid development of evidence-based preventive measures for occupational accidents.

## I. Introduction

The number of injuries and fatalities from occupational accidents requiring four or more days of leave remains high, with “falls” being the most common type of incident.^1^ The health/hygiene and retail industries, in particular, have a high incidence of fall-related accidents, making the formulation of effective countermeasures an urgent priority.

Our previous research analyzed fall accidents in these two sectors using the Ministry of Health, Labour and Welfare’s industrial accident database. We found that 35.9% of accidents occurred outdoors, with slipping on snow and ice being a primary cause, especially during winter.^2^ This insight was gained through a meticulous process where experts reviewed the text data describing the accident circumstances and carefully classified items such as “accident location” and “cause of accident.” However, this manual classification process, while standard in various studies,^3-^□ is extremely time-consuming and labor-intensive. Previous studies using the same Japanese industrial accident database have employed methods like rule-based text searches for classification,□ ^-^□ but these approaches are limited in their applicability to large datasets, speed of analysis, and ability to handle the diverse expressions found in free-text descriptions.

The application of machine learning, a type of AI, could solve this problem. However, traditional methods require task-specific pre-training, □ which in turn necessitates the manual preparation of a large volume of labeled training data. This reliance on manual labor for data preparation creates a dilemma, as it does not lead to a fundamental reduction in workload.

Recent advancements in Large Language Models (LLMs) are changing this landscape. LLMs are pre-trained on an extremely broad range of knowledge and are not specialized for specific tasks. Therefore, we hypothesized that by directly leveraging the general-purpose language capabilities of LLMs, we could automate and streamline the analysis of occupational accidents, bypassing the task-specific model training process required by conventional machine learning. While there have been reports of LLMs being used for labeling in the medical field,^1^ □ no such reports were found in the field of industrial hygiene.

## II. Methods

### Study Design

This is an accuracy evaluation study that compares the results of automatic classification by LLMs with the results of manual classification by experts.

### Analysis Data

The data for this analysis was drawn from the 2021 Survey on Industrial Accidents database,^11^ previously used in a prior study. It consists of data on injuries requiring four or more days of leave in the health/hygiene and wholesale/retail industries. This data is a random sample of approximately one-quarter of the worker injury and illness reports submitted by employers and is publicly available as anonymized open data. Each case includes text data describing the specific circumstances of the accident (“Accident Description”). The 2021 database contains 29,605 cases. From these, we selected 2,690 cases classified as falls in the specified industries. After excluding 71 cases where manual review determined that a fall did not occur or it was unclear, 2,619 cases remained as the final study subjects.

### 1. Creation of Ground Truth Data

The “ground truth” data, which served as the benchmark for evaluating the AI’s classification accuracy in this study, was derived from the manual classification results created by experts in our previous research.^2^ The process was as follows: First, a physical therapist with expertise in the rehabilitation of fall-related injuries read through the accident description text for all 2,619 cases one by one. They then created a draft classification for six categories: “Within/Outside Business Premises,” “Accident Location,” “In a Vehicle,” “Cause of Accident,” “Causal Substance,” and “Injured Body Part.” Next, an occupational physician with expertise in occupational health reviewed this draft classification for cases that were flagged as unclear or difficult to judge in the initial assessment. This entire process took several weeks to complete.

### 2. Automatic Classification by Large Language Models (LLMs)

To automate the manual classification described above, this study utilized Large Language Models (LLMs), a type of AI. We selected four models with varying performance from the well-known GPT series developed by OpenAI: GPT-4.1, GPT-4.1 mini, GPT-4o mini, and o4-mini, to compare their performance and cost. The processing was conducted in June 2025, using the latest versions of each model available at that time.

The processing was implemented on Google Colaboratory,^12^ a cloud-based execution environment, using Python (3.11) and the OpenAI library (1.84.0).^13^ To efficiently process thousands of cases, we utilized the Batch API feature. This method allows multiple requests to be sent as a single job, which is then processed asynchronously on the server side. While it can take up to 24 hours, it is available at half the cost of synchronous processing, making it suitable for executing a large number of mutually independent tasks like ours. However, we observed that batch jobs would occasionally stall. To mitigate this, we designed a system that split the tasks into chunks of 100, submitted them all at once, monitored their status at regular intervals, and was able to cancel and recreate any tasks that were identified as stalled.

For the seven classification items under investigation, we created a specific prompt (instruction) for each and executed them as individual tasks in the batch process. The original prompts given to the AI are shown in Table 1.

**Table 1.**
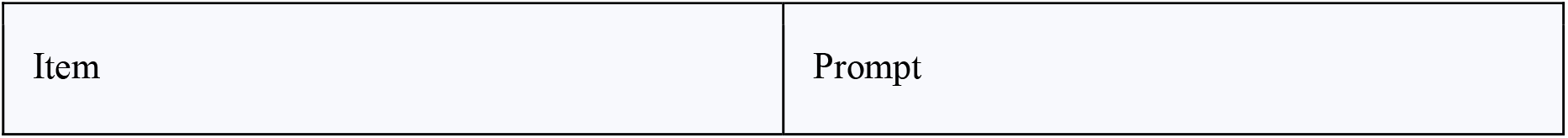
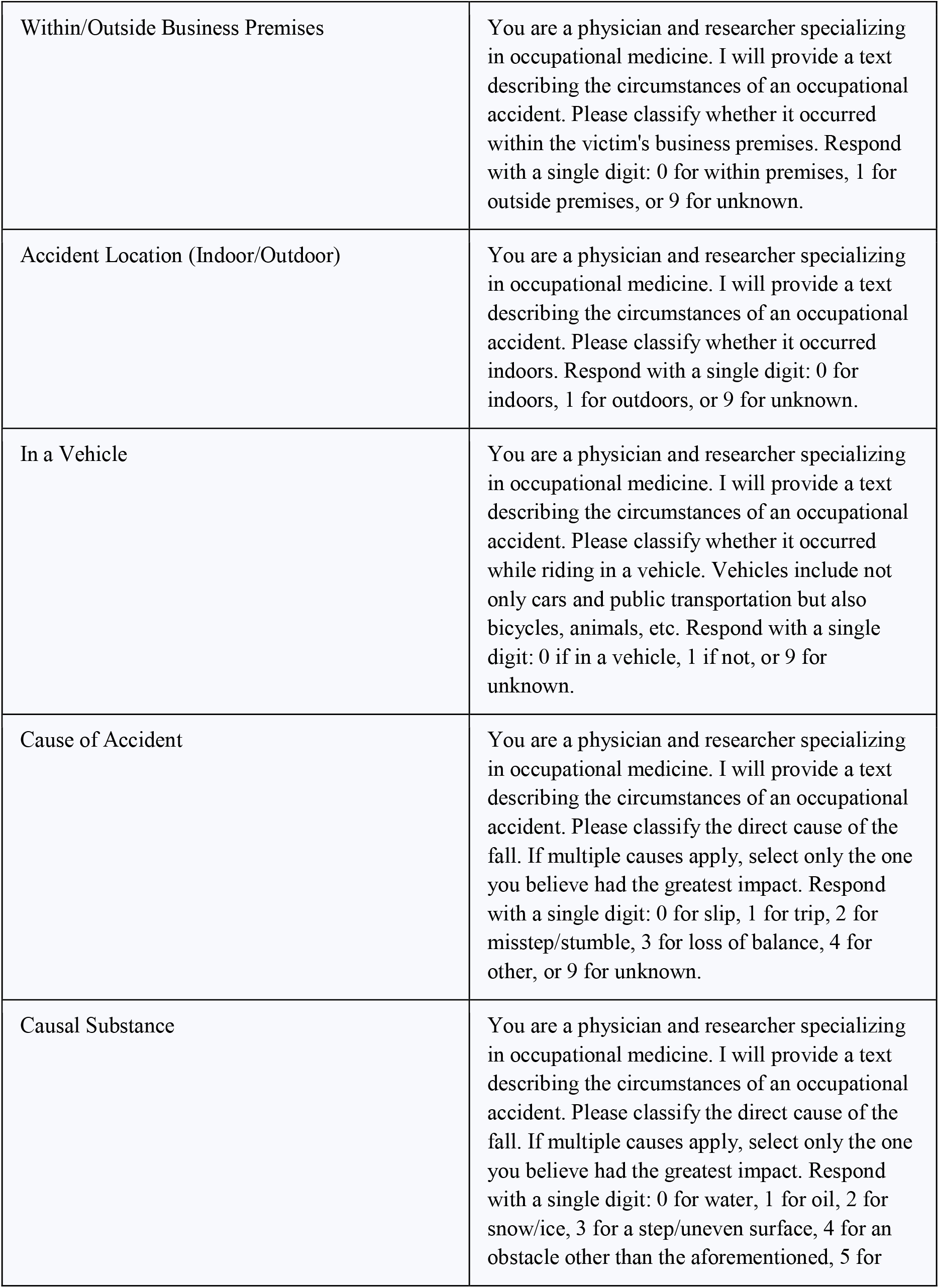
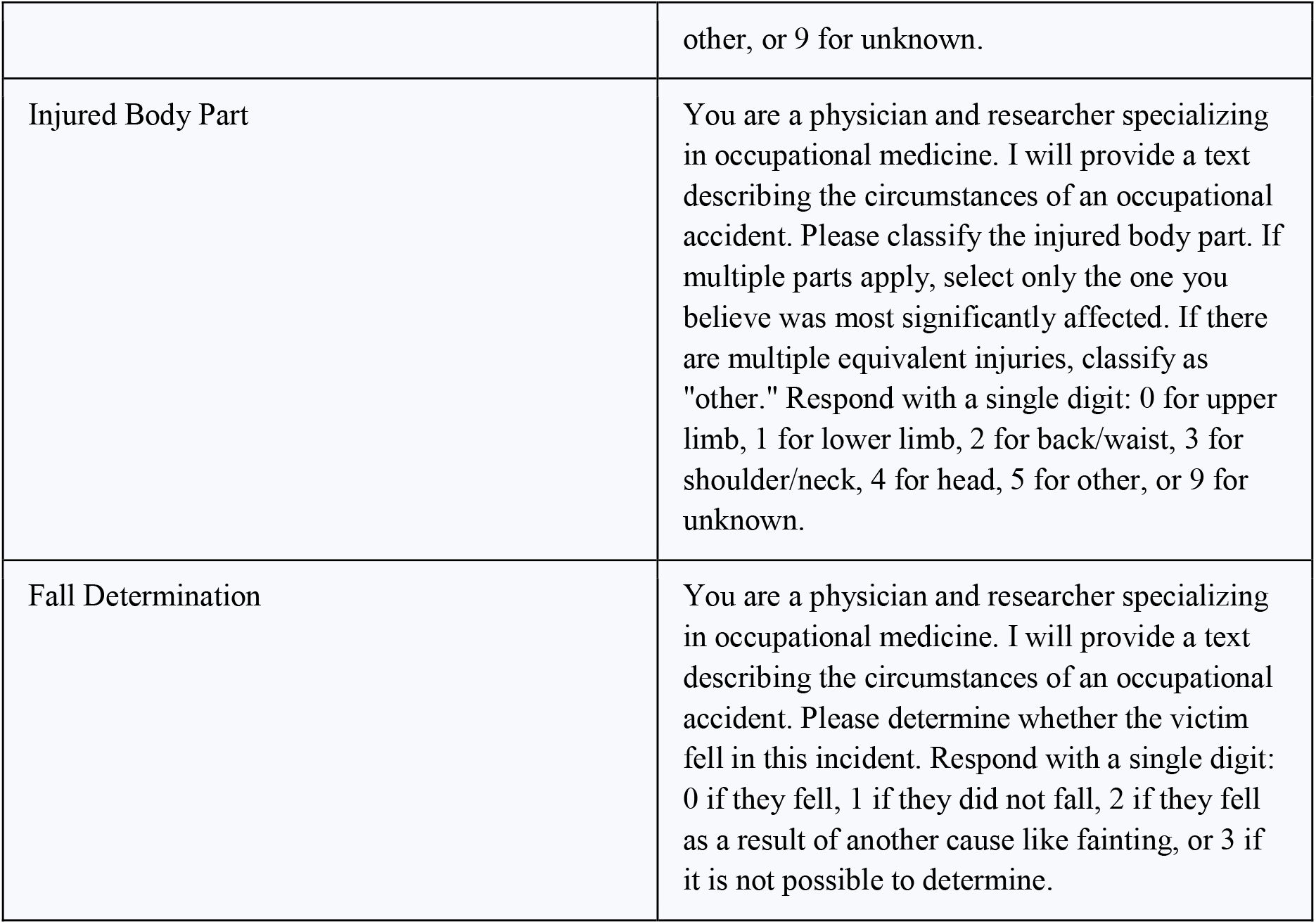
Original Prompts Input to LLM for Each Classification Item.

| Item | Prompt |
| --- | --- |
| Within/Outside Business Premises | You are a physician and researcher specializing in occupational medicine. I will provide a text describing the circumstances of an occupational accident. Please classify whether it occurred within the victim's business premises. Respond with a single digit: 0 for within premises, 1 for outside premises, or 9 for unknown. |
| Accident Location (Indoor/Outdoor) | You are a physician and researcher specializing in occupational medicine. I will provide a text describing the circumstances of an occupational accident. Please classify whether it occurred indoors. Respond with a single digit: 0 for indoors, 1 for outdoors, or 9 for unknown. |
| In a Vehicle | You are a physician and researcher specializing in occupational medicine. I will provide a text describing the circumstances of an occupational accident. Please classify whether it occurred while riding in a vehicle. Vehicles include not only cars and public transportation but also bicycles, animals, etc. Respond with a single digit: 0 if in a vehicle, 1 if not, or 9 for unknown. |
| Cause of Accident | You are a physician and researcher specializing in occupational medicine. I will provide a text describing the circumstances of an occupational accident. Please classify the direct cause of the fall. If multiple causes apply, select only the one you believe had the greatest impact. Respond with a single digit: 0 for slip, 1 for trip, 2 for misstep/stumble, 3 for loss of balance, 4 for other, or 9 for unknown. |
| Causal Substance | You are a physician and researcher specializing in occupational medicine. I will provide a text describing the circumstances of an occupational accident. Please classify the direct cause of the fall. If multiple causes apply, select only the one you believe had the greatest impact. Respond with a single digit: 0 for water, 1 for oil, 2 for snow/ice, 3 for a step/uneven surface, 4 for an obstacle other than the aforementioned, 5 for |
|  | other, or 9 for unknown. |
| Injured Body Part | You are a physician and researcher specializing in occupational medicine. I will provide a text describing the circumstances of an occupational accident. Please classify the injured body part. If multiple parts apply, select only the one you believe was most significantly affected. If there are multiple equivalent injuries, classify as "other." Respond with a single digit: 0 for upper limb, 1 for lower limb, 2 for back/waist, 3 for shoulder/neck, 4 for head, 5 for other, or 9 for unknown. |
| Fall Determination | You are a physician and researcher specializing in occupational medicine. I will provide a text describing the circumstances of an occupational accident. Please determine whether the victim fell in this incident. Respond with a single digit: 0 if they fell, 1 if they did not fall, 2 if they fell as a result of another cause like fainting, or 3 if it is not possible to determine. |

Furthermore, the parameters (settings) for the API calls are crucial for ensuring the reliability of the results (Table 2). In this study, we set the temperature, which controls the “creativity” of the AI, to 0. This minimizes random elements, ensures that the AI generally returns the same answer for the same input, and thus secures the reproducibility of the research.

**Table 2.** Main API Parameters.

| Parameter | Description | Value |
| --- | --- | --- |
| model | The name of the LLM used. | "GPT-4.1, GPT-4.1 mini, GPT-4o-mini, o4-mini" |
| temperature | Controls the randomness (creativity) of the output. Set to 0 to ensure reproducibility. | 0 |
| max_tokens | The maximum length of the generated response. Set to a short value as a single-digit | 10 |
|  | response is expected. |  |
| Note: The reasoning model o4-mini does not support temperature and max_tokens, so these were not set. |  |  |

### 3. Accuracy Evaluation

The results classified by each LLM were compared with the expert’s manual classification results (ground truth data) to evaluate their level of agreement. We adopted commonly used metrics for evaluating the performance of classification models. Since our classification task is a multi-class classification with three or more categories (e.g., “Accident Location” has indoors, outdoors, and unknown), each metric was calculated per category and then averaged.

- Accuracy: The proportion of correctly classified data out of the total data. Accuracy = (Number of Correctly Classified Data) / (Total Number of Data)
- Precision: Of the items predicted as a certain category (e.g., “outdoors”), the proportion that was actually correct. It indicates how well false positives were suppressed. Precision = TP / (TP + FP) (TP: True Positive, FP: False Positive)
- Recall: Of the items that are actually in a certain category (e.g., “outdoors”), the proportion that the AI was able to identify. It indicates how few false negatives there were. Recall = TP / (TP + FN) (FN: False Negative)
- F1-Score: The harmonic mean of precision and recall. It is used when a balanced evaluation of both metrics is desired. A value closer to 1 indicates better performance. F1-Score = 2 * (Precision * Recall) / (Precision + Recall)
- Cohen’s Kappa Score: Unlike simple accuracy, this metric calculates the agreement between the model’s predictions and the true labels after accounting for agreement that could occur by chance. This makes it a more robust measure of performance, especially for imbalanced datasets.

The precision, recall, and F1-score presented in this paper are weighted averages, where the metric for each category is weighted by the number of data points (support) in that category. This accounts for class imbalance and provides a more realistic evaluation of the overall dataset performance. All metrics were calculated using scikit-learn (1.6.1).

### 4. Ethical Considerations

This study exclusively used the publicly available, fully anonymized Survey on Industrial Accidents database provided by the Ministry of Health, Labour and Welfare. Therefore, it falls outside the scope of the “Ethical Guidelines for Life Science and Medical Research Involving Human Subjects” and did not require approval from an ethics review committee.

## III. Results

### 1. Classification Accuracy

We automatically classified 2,619 fall accident cases using four LLMs and evaluated their accuracy against the manual classification results as the ground truth. This paper reports the results for four representative classification items. The results are shown in Table 3. Notably, in one case, GPT-4.1 mini provided a text response instead of the instructed number; this was manually reclassified as unknown.

**Table 3.** List of Evaluation Metrics for Each Item and Model.

|  | Accuracy | Precision | Recall | F1-Score | Kappa |
| --- | --- | --- | --- | --- | --- |
| Indoor/Outdoor |  |  |  |  |  |
| GPT-4o mini | 0.911 | 0.892 | 0.911 | 0.901 | 0.810 |
| GPT-4.1 mini | 0.921 | 0.914 | 0.921 | 0.917 | 0.835 |
| GPT-4.1 | 0.923 | 0.916 | 0.923 | 0.919 | 0.838 |
| o4-mini | 0.913 | 0.929 | 0.913 | 0.920 | 0.824 |
| Cause of Accident |  |  |  |  |  |
| GPT-4o mini | 0.796 | 0.758 | 0.796 | 0.751 | 0.715 |
| GPT-4.1 mini | 0.807 | 0.805 | 0.807 | 0.770 | 0.734 |
| GPT-4.1 | 0.789 | 0.721 | 0.789 | 0.732 | 0.710 |
| o4-mini | 0.818 | 0.846 | 0.818 | 0.784 | 0.749 |
| Causal Substance |  |  |  |  |  |
| GPT-4o mini | 0.371 | 0.601 | 0.371 | 0.332 | 0.283 |
| GPT-4.1 mini | 0.599 | 0.671 | 0.599 | 0.535 | 0.510 |
| GPT-4.1 | 0.638 | 0.732 | 0.638 | 0.583 | 0.559 |
| o4-mini | 0.728 | 0.747 | 0.728 | 0.709 | 0.662 |
| Injured Body Part |  |  |  |  |  |
| GPT-4o mini | 0.798 | 0.847 | 0.798 | 0.790 | 0.699 |
| GPT-4.1 mini | 0.849 | 0.872 | 0.849 | 0.849 | 0.774 |
| GPT-4.1 | 0.840 | 0.868 | 0.840 | 0.845 | 0.764 |
| o4-mini | 0.843 | 0.878 | 0.843 | 0.850 | 0.771 |

Overall, the oldest model, GPT-4o mini, performed the poorest, with its accuracy for “causal substance” dropping to just 37%. In contrast, newer models achieved accuracies of 60-73% for the same item. For the other items, accuracy was generally between 70% and 90%, with kappa scores around 0.7 to 0.84.

For the “causal substance” category, which had the lowest accuracy, the confusion matrices for the worst-performing model (GPT-4o mini) and the best-performing model (o4-mini) are shown in Figure 1 and Figure 2, respectively.

**Figure 1.**
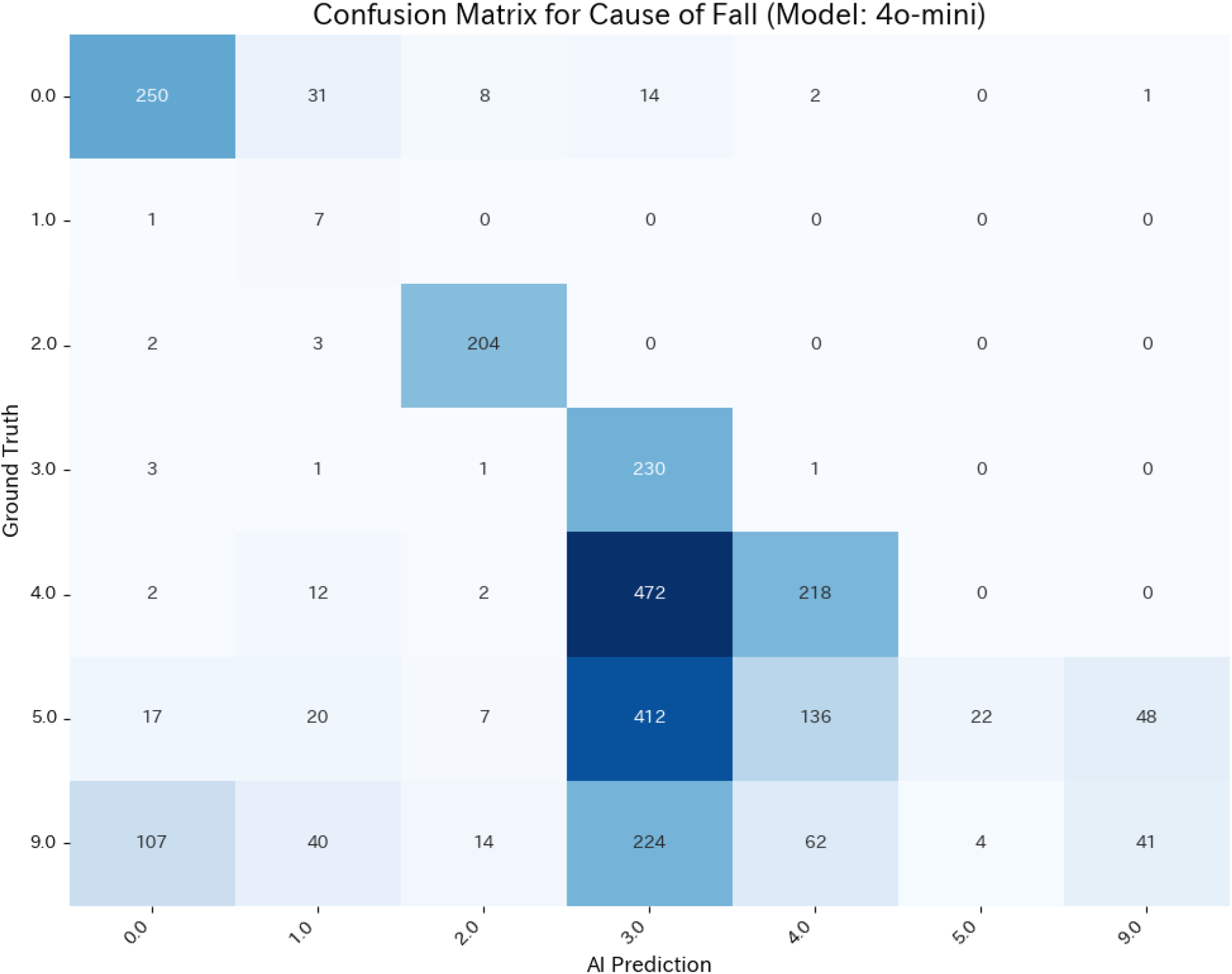
Confusion Matrix for Causal Substance of Fall (Model: GPT-4o mini)

**Figure 2.**
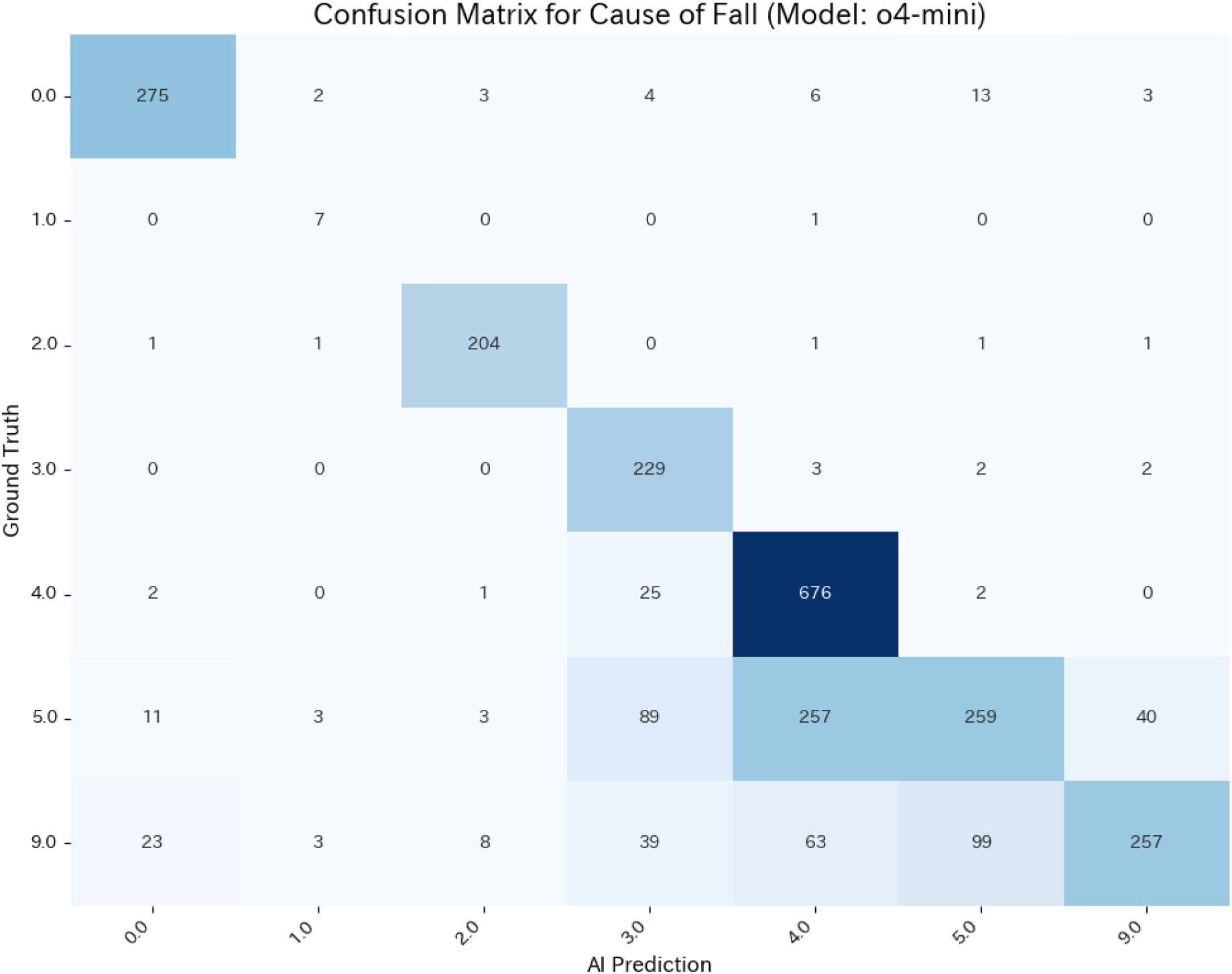
Confusion Matrix for Causal Substance of Fall (Model: o4-mini)

### 2. Processing Time and Cost

The time and estimated cost for processing all 2,619 cases using the Batch API for the three models are shown in Table 4. Even with the most expensive model, o4-mini, the processing was completed in about 90 minutes for approximately $11. The lightweight model, GPT-4.1 mini, took about 24 minutes and cost $0.75.

**Table 4.** Processing Time and Estimated Cost for Batch Processing with Each Model.

| Model | Price (USD) | Time* (h:m:s) |
| --- | --- | --- |
| GPT-4o mini | \$0.28 | 2:53:55 |
| GPT-4.1 mini | \$0.75 | 0:24:04 |
| GPT-4.1 | \$3.73 | 0:19:19 |
| o4-mini | \$10.58 | 1:27:45 |

\*When using the Batch API, jobs are executed within 24 hours based on resource availability, so processing times may vary significantly.

## IV. Discussion

This study automated the classification of occupational accident text data using LLMs and verified its accuracy and practicality. The results indicate that LLMs can replicate the manual classification work that took experts several weeks, doing so with high speed and accuracy.

### Advantages Over Traditional Machine Learning

The LLM-based approach adopted in this study holds a decisive advantage over traditional machine learning models like deep learning: it eliminates the need for task-specific training.

When building a text classification model with conventional methods, one must first manually prepare thousands of “training data” samples, which are then split into “training sets” and “evaluation sets.” The model learns specific classification patterns from the training set and is then tested on the unseen evaluation data. This process of creating and training with labeled data is itself a specialized and time-consuming task, posing a major obstacle to automation. For a dataset of several thousand items like ours, the manual labeling for the training data would essentially complete the classification for the entire set, limiting the utility of machine learning.

In contrast, the method used in this study achieves high-accuracy classification with only simple prompts, without any training. Unlike humans, LLMs are not susceptible to fatigue or inconsistencies in judgment, allowing for stable and high-quality classification. This “training-free” characteristic dramatically reduces the initial cost and preparation time for analysis, making advanced text analysis accessible to more researchers and practitioners.

### Practicality of Using the Batch API

As shown in Table 4, even with the most expensive model, o4-mini, the classification of 2,619 cases was completed in about 90 minutes for approximately $11. This represents a significant improvement in efficiency compared to the time and labor costs of manual expert classification. This result suggests that the combination of LLMs and the Batch API is a viable method for data analysis in the occupational safety and health field. This efficiency opens the door to larger-scale and more frequent analyses that were previously prohibitive due to cost and time constraints. For example, it could enable monthly analysis of national occupational accident data to detect early signs of new risks or rapid, detailed factor analysis for specific industries or tasks. The worker injury reports that form the basis of this database, originally prepared and submitted by hand, are slated to be submitted electronically as a general rule from January 2025.^1^ □ This transition is expected to further accelerate the speed of analysis.

### On Classification Accuracy

Among the models used, GPT-4o mini was the oldest and performed the worst. Its successor, GPT-4.1, and its lightweight version, GPT-4.1 mini, achieved accuracies of 70-80% for items other than “causal substance,” with kappa coefficients generally exceeding 0.7. According to Landis & Koch’s criteria,^1^ □ the agreement for all items would be rated as “Substantial” or higher, with some even classified as “Almost Perfect.” This indicates that LLM-based classification is comparable to that of experts.

The lower accuracy for “causal substance” seems to stem partly from the inherent ambiguity of the categories, with significant confusion observed between “other obstacles,” “other,” and “unknown.” The definitions for these categories were not explicitly set and were left to the discretion of the human raters, which likely posed a difficult task for the AI. However, the o4-mini model,^1^ □ which is specialized for reasoning through thought processes, achieved a kappa coefficient over 0.6, indicating a respectable level of accuracy. It is presumed that its reasoning process led to judgments more aligned with human intuition. A study in the public health domain that attempted to annotate social media data using GPT-4 Turbo reported lower LLM accuracy for tasks requiring contextual understanding,^1^ □ which aligns with our findings. However, our study also tested newer models, and as noted, these results suggest that improvements are being made in this area.

These results suggest that AI-based classification possesses a certain degree of accuracy. Depending on the application, required accuracy, and speed, one could consider using generative AI for full classification, as one of the classifiers in a team, or as an auxiliary tool. It may also be useful to perform a preliminary AI-based classification even before manual classification to check whether the classification criteria and content are clearly defined.

### Limitations of this Study

This study has several limitations. First, the validation is limited to a specific dataset, and whether similar performance can be achieved for other industries or types of accidents requires separate verification. Second, the performance of LLMs depends on the quality of the input text data. The data used in this study was created by various businesses, with some descriptions being detailed and others not. A lack of detail could lead to more guesswork and reduced accuracy. Our evaluation was conducted on a full year’s worth of actual data, demonstrating a certain level of accuracy even with this variability. Third, the prompts used in this study are just one example, and accuracy could potentially be improved with further refinement (e.g., few-shot learning). Nevertheless, we believe it is significant that a certain level of accuracy was achievable even with simple prompts.

## V. Conclusion

This study demonstrates that in the classification of text data from occupational accident reports, LLMs can serve as a high-accuracy substitute for manual expert labor. Furthermore, by utilizing the Batch API, large-scale data analysis can be performed efficiently and at a low cost. The application of this method facilitates large-scale, multi-faceted accident analysis, which has been difficult due to time and human resource costs, and can contribute to the rapid formulation of evidence-based occupational accident prevention measures. The findings of this research represent an important step in advancing occupational safety and health intelligence by leveraging AI technology that does not require task-specific model training.

## Data Availability

All data produced in the present study are available upon reasonable request to the corresponding author.

## Author Contribution

H.A. and R.M. conceived and designed the study. H.A., R.M., and S.Y. were responsible for the data collection and investigation. H.A. performed the statistical analysis and data visualization. H.A. was also responsible for project administration and wrote the first draft of the manuscript. A.O. supervised the project. All authors participated in the revision of the manuscript and approved the final version for submission.

## Ethical Statement

Ethical review and approval were not required for this study as it was based solely on publicly available, anonymized data, in accordance with the local legislation and institutional requirements.

## References

1. Ministry of Health, Labour and Welfare, Japan. Occupational Accident Statistics for 2023. https://www.mhlw.go.jp/stf/newpage_40395.html (in Japanese)

2. Matsugaki R, Yamakawa S, Ando H, Ogami A. Same-Level Fall Injuries among Healthcare and Retail Workers: Focus on Outdoor Incidents. SANGYO EISEIGAKU ZASSHI. 2025(in press, in Japanese)

3. Tsai TY, Lin JF, Tu YK, et al. Validation of ICD-10-CM Diagnostic Codes for Identifying Patients with ST-Elevation and Non-ST-Elevation Myocardial Infarction in a National Health Insurance Claims Database. Clin Epidemiol. 2023;15:1027–1039. doi:10.2147/clep.S431231

4. So L, Evans D, Quan H. ICD-10 coding algorithms for defining comorbidities of acute myocardial infarction. BMC Health Serv Res. 2006;6(1) doi:10.1186/1472-6963-6-161

5. Nakai M, Iwanaga Y, Sumita Y, et al. Validation of Acute Myocardial Infarction and Heart Failure Diagnoses in Hospitalized Patients With the Nationwide Claim-Based JROAD-DPC Database. Circulation Reports. 2021;3(3):131–136. doi:10.1253/circrep.cr-21-0004

6. Wasaki N, Takahashi A. Characteristics of Occupational Accidents Caused by Inattentiveness. Journal of Occupational Safety and Health. 2024;17(2):93–104. doi:10.2486/josh.JOSH-2023-0016-GE (in Japanese)

7. Sugama A. Present Situation of Falls from Step Ladders and Future Perspectives on Preventative Countermeasures. Journal of Occupational Safety and Health. 2017;10(1):55–58. doi:10.2486/josh.JOSH-2016-0010-SHI (in Japanese)

8. Hayashi C, Ogata S, Toyoda H, et al. Risk factors for fracture by same-level falls among workers across sectors: a cross-sectional study of national open database of the occupational injuries in Japan. Public Health. 2023;217:196–204. doi:10.1016/j.puhe.2023.02.003

9. Lu H, Ehwerhemuepha L, Rakovski C. A comparative study on deep learning models for text classification of unstructured medical notes with various levels of class imbalance. BMC Med Res Methodol. 2022;22(1) doi:10.1186/s12874-022-01665-y

10. Balch JA, Desaraju SS, Nolan VJ, et al. Language Models for Multilabel Document Classification of Surgical Concepts in Exploratory Laparotomy Operative Notes: Algorithm Development Study. JMIR Medical Informatics. 2025;13:e71176–e71176. doi:10.2196/71176

11. Ministry of Health, Labour and Welfare, Japan. Database of Serious Occupational Accidents (Fatalities and Cases Involving Four or More Days of Leave). https://anzeninfo.mhlw.go.jp/anzen_pgm/SHISYO_FND.html (in Japanese)

12. Google LLC. Google Colaboratory. https://colab.google/

13. OpenAI Inc. OpenAI Platform. https://platform.openai.com/docs/overview

14. Ministry of Health, Labour and Welfare, Japan. The reporting requirements for the Report of Worker Death, Injury, or Illness will be revised, and electronic submission will become mandatory (effective January 1, 2025). https://www.mhlw.go.jp/stf/seisakunitsuite/bunya/koyou_roudou/roudoukijun/denshishinsei_00002.html (in Japanese)

15. Landis JR, Koch GG. The measurement of observer agreement for categorical data. Biometrics. Mar 1977;33(1):159–74.

16. OpenAI Inc. OpenAI o3 and o4-mini System Card. https://cdn.openai.com/pdf/2221c875-02dc-4789-800b-e7758f3722c1/o3-and-o4-mini-system-card.pdf

17. Kazari K, Chen Y, Shakeri Z. Scaling Public Health Text Annotation: Zero-Shot Learning vs. Crowdsourcing for Improved Efficiency and Labeling Accuracy. arXiv. 2025; doi:10.48550/arxiv.2502.06150

